## Supplemental Table S1 for "Recessive pathogenic variants in *MCAT* cause combined oxidative phosphorylation deficiency"

**Supplemental Table S1. Absence of Heterozygosity (AOH) identified on chromosomal microarray**

| AOH Region [GRCh37] | Size (bp) | No. of genes |
|---|---:|---:|
| 1p32.3p32.2(52407782_57646055) | 5,238,274 | 107 |
| 1q41q42.12(219978259_226762648) | 6,784,390 | 109 |
| 2q23.3q24.1(150509332_155344473) | 4,835,142 | 37 |
| 9p24.1p23(7068436_9752968) | 2,684,533 | 11 |
| 9q21.31q21.32(81625681_84241442) | 2,615,762 | 23 |
| 22q12.3q13.31(36074596_46172372) | 10,097,777 | 292 |
