## Supplemental Table S2 for "Recessive pathogenic variants in *MCAT* cause combined oxidative phosphorylation deficiency"

Supplemental Table S2.  Metabolomics data.

| Sample | P1-1 | P1-2 | P1-3 | C5-1 | C5-2 | C5-3 | C4-1 | C4-2 | C4-3 | C3-1 | C3-2 | C3-3 | C1-1 | C1-2 | C1-3 | C2-1 | C2-2 | C2-3 |
|---|---|---|---|---|---|---|---|---|---|---|---|---|---|---|---|---|---|---|
| Label | Proband | Proband | Proband | Control | Control | Control | Control | Control | Control | Control | Control | Control | Control | Control | Control | Control | Control | Control |
| Acetyl-CoA | 8.28E-05 | 8.72E-05 | 1.28E-04 | 7.53E-05 | 1.28E-04 | 5.14E-05 | 9.02E-07 | 3.98E-05 | 2.41E-05 | 3.01E-05 | 7.44E-05 | 5.79E-05 | 2.24E-04 | 1.54E-04 | 2.23E-04 | 2.08E-04 | 2.6E-04 | 1.52E-04 |
| ASPARTATE | 1.1E-02 | 1.37E-02 | 1.79E-02 | 1.22E-02 | 1.77E-02 | 1.15E-02 | 9.78E-03 | 6.18E-03 | 6.26E-03 | 8.16E-03 | 1.22E-02 | 1.22E-02 | 2.31E-02 | 1.75E-02 | 1.55E-02 | 2.56E-02 | 2.32E-02 | 2.31E-02 |
| CITRATE/ ISOCITRATE | 3.92E-02 | 3.41E-02 | 6.34E-02 | 4.19E-02 | 5.26E-02 | 3.88E-02 | 2.82E-02 | 1.94E-02 | 3.03E-02 | 3.04E-02 | 3.56E-02 | 6.19E-02 | 1.34E-01 | 9.11E-02 | 7.47E-02 | 9.89E-02 | 9.02E-02 | 6.97E-02 |
| FUMARATE | 8.25E-04 | 8.08E-04 | 1.2E-03 | 9.14E-04 | 1.05E-03 | 7.39E-04 | 5.4E-04 | 3.38E-04 | 4.29E-04 | 5.12E-04 | 6.88E-04 | 1.06E-03 | 1.79E-03 | 1.14E-03 | 9.27E-04 | 1.74E-03 | 1.64E-03 | 1.31E-03 |
| LACTATE | 3.5E-03 | 4.13E-03 | 5.02E-03 | 2.83E-03 | 2.52E-03 | 2.87E-03 | 3.16E-03 | 2.07E-03 | 2.98E-03 | 2.57E-03 | 2.67E-03 | 2.56E-03 | 4.85E-03 | 2.49E-03 | 2.75E-03 | 2.72E-03 | 2.77E-03 | 2.94E-03 |
| MALATE | 4.79E-03 | 4.82E-03 | 7.37E-03 | 5.23E-03 | 6.28E-03 | 4.6E-03 | 3.15E-03 | 1.95E-03 | 2.37E-03 | 2.7E-03 | 3.65E-03 | 5.66E-03 | 1.05E-02 | 6.58E-03 | 5.56E-03 | 1.05E-02 | 9.41E-03 | 7.79E-03 |
| OXOGLUTARATE | 1.19E-04 | 9.83E-05 | 1.57E-04 | 1.42E-04 | 1.54E-04 | 1.16E-04 | 1.24E-04 | 5.86E-05 | 8.79E-05 | 9.69E-05 | 7.16E-05 | 1.31E-04 | 2.25E-04 | 1.58E-04 | 1.32E-04 | 2.14E-04 | 2.29E-04 | 1.61E-04 |
| SUCCINATE | 2.06E-04 | 2.15E-04 | 3.31E-04 | 1.87E-04 | 1.7E-04 | 1.78E-04 | 1.89E-04 | 1.04E-04 | 1.65E-04 | 1.46E-04 | 2.03E-04 | 2.7E-04 | 3.68E-04 | 2.03E-04 | 2.3E-04 | 2.63E-04 | 2.29E-04 | 2.41E-04 |
| PYRUVATE | 2.15E-05 | 1.2E-06 | 5.89E-05 | 2.93E-05 | 3.16E-05 | 5.33E-05 | 3.33E-05 | 2.62E-05 | 3.78E-05 | 3.82E-05 | 4.07E-05 | 3.33E-05 | 1.53E-06 | 2.21E-05 | 2.97E-05 | 1.23E-06 | 9.77E-07 | 1.86E-05 |
| 10-HYDROXYDECANOATE | 7.57E-05 | 8.41E-05 | 9.07E-05 | 6.6E-05 | 5.89E-05 | 4.97E-05 | 8.2E-05 | 6.03E-05 | 6.88E-05 | 8.26E-05 | 7.11E-05 | 4.34E-05 | 1.18E-04 | 7.42E-05 | 4.68E-05 | 8.52E-05 | 5.3E-05 | 8.62E-05 |
| 2-DEOXY-D-GLUCOSE | 1.41E-03 | 1.37E-03 | 3.91E-03 | 2.95E-04 | 2.34E-03 | 8.84E-04 | 1.81E-03 | 3.56E-05 | 4.78E-04 | 2.75E-03 | 3.28E-04 | 2.1E-03 | 4.59E-03 | 3.79E-04 | 1.25E-04 | 1.73E-03 | 1.47E-03 | 6.44E-04 |
| 2-HYDROXYBUTYRATE | 2.3E-05 | 3.23E-05 | 3.98E-05 | 2.15E-05 | 2E-05 | 2.32E-05 | 2.72E-05 | 2.18E-05 | 2.69E-05 | 2.4E-05 | 2.5E-05 | 1.87E-05 | 4.35E-05 | 2.07E-05 | 1.88E-05 | 2.87E-05 | 2.01E-05 | 2.2E-05 |
| 3-HYDROXYMETHYLGLUTARATE | 6.38E-04 | 8.59E-04 | 1.03E-03 | 5.26E-04 | 6.68E-04 | 4.91E-04 | 6.29E-04 | 4.66E-04 | 5.09E-04 | 5.26E-04 | 7.3E-04 | 6.88E-04 | 1.18E-03 | 7.22E-04 | 6.42E-04 | 1.12E-03 | 9.11E-04 | 9.55E-04 |
| 3-METHYL-2-OXOVALERATE | 2.47E-03 | 3.81E-03 | 3.23E-03 | 1.52E-03 | 1.31E-03 | 1.49E-03 | 1.78E-03 | 1.59E-03 | 2.14E-03 | 2.23E-03 | 2.45E-03 | 1.48E-03 | 3.07E-03 | 1.46E-03 | 1.5E-03 | 2.17E-03 | 1.89E-03 | 2.26E-03 |
| 3-Methylglutarylcarnitine | 2.09E-05 | 2.03E-05 | 1.1E-06 | 7.46E-07 | 1.03E-06 | 1.61E-05 | 1.29E-05 | 2.13E-05 | 3.1E-05 | 9.7E-07 | 3.01E-05 | 3.94E-05 | 1.53E-06 | 1.01E-06 | 1.36E-05 | 2.76E-05 | 2.07E-05 | 1.93E-05 |
| 4-AMINOBENZOATE | 8.25E-07 | 9.94E-05 | 9.78E-05 | 6.35E-05 | 1.03E-06 | 7.4E-05 | 7E-05 | 6.49E-05 | 9.56E-07 | 8.49E-05 | 7.63E-05 | 9.06E-05 | 1.32E-04 | 9.43E-05 | 7.89E-05 | 1.07E-04 | 9.77E-07 | 1.01E-04 |
| 4-IMIDAZOLEACETATE | 2.95E-04 | 1.69E-04 | 6.91E-05 | 3.29E-04 | 1.55E-04 | 2.14E-04 | 1.18E-04 | 2.19E-04 | 1.91E-04 | 1.25E-04 | 1.61E-04 | 9.11E-05 | 9.48E-05 | 1.48E-04 | 3.36E-04 | 1.67E-04 | 2.13E-04 | 2.27E-04 |
| 4-PYRIDOXATE | 4.01E-04 | 5.25E-04 | 2.82E-05 | 4.34E-04 | 3.35E-04 | 4.45E-04 | 5.06E-04 | 4.53E-04 | 6.42E-04 | 4.78E-04 | 4.82E-04 | 3.74E-04 | 7E-04 | 3.12E-04 | 3.29E-04 | 2.97E-04 | 2.84E-04 | 3.37E-04 |
| 4-QUINOLINECARBOXYLATE | 1.42E-05 | 6.1E-05 | 6.28E-05 | 2.06E-05 | 5.11E-05 | 3.79E-05 | 2.47E-05 | 1.6E-05 | 5.46E-05 | 4.98E-05 | 5.61E-05 | 3.63E-05 | 7.64E-05 | 7.59E-05 | 3.63E-05 | 1.08E-05 | 9.35E-06 | 2.91E-05 |
| 5-AMINOLEVULINATE | 8E-04 | 1.47E-03 | 7.43E-04 | 3.03E-04 | 1.5E-04 | 4.63E-05 | 4.47E-04 | 5.31E-04 | 1E-03 | 2.78E-04 | 7.88E-04 | 5.84E-04 | 1.53E-03 | 2.41E-04 | 1.83E-04 | 1.23E-03 | 5.69E-04 | 7.77E-04 |
| 5-AMINOPENTANOATE | 6.85E-05 | 1.57E-04 | 8.87E-05 | 4.58E-05 | 6.93E-05 | 4.66E-05 | 6.3E-05 | 6.08E-05 | 8.55E-05 | 5E-05 | 1.07E-04 | 7.6E-05 | 1.58E-04 | 5.19E-05 | 4.13E-05 | 1.14E-04 | 7.94E-05 | 1.04E-04 |
| 5-Methylcytidine | 1.08E-04 | 2.02E-04 | 1.79E-04 | 1.52E-04 | 1.41E-04 | 1.53E-04 | 1.77E-04 | 1.54E-04 | 1.67E-04 | 1.01E-04 | 9.13E-07 | 1.26E-04 | 1.53E-06 | 1.01E-06 | 1.74E-04 | 1.61E-04 | 1.25E-04 | 1.01E-06 |
| 6-HYDROXYDOPAMINE | 1.27E-01 | 8.59E-02 | 7.91E-02 | 1.23E-01 | 1.06E-01 | 1.33E-01 | 1.25E-01 | 1.33E-01 | 1.16E-01 | 1.26E-01 | 9.77E-02 | 1.12E-01 | 4.29E-02 | 9.88E-02 | 1.12E-01 | 8.57E-02 | 9.95E-02 | 9.37E-02 |
| 8-HYDROXY-DEOXYGUANOSINE | 1.14E-04 | 1.64E-04 | 1.6E-04 | 6.95E-05 | 1.02E-04 | 8.44E-05 | 8.21E-05 | 8.9E-05 | 9.49E-05 | 9.77E-05 | 1.37E-04 | 9.75E-05 | 1.18E-04 | 1.08E-04 | 7.62E-05 | 1.08E-04 | 9.92E-05 | 1.02E-04 |
| ADENINE | 3.16E-03 | 4.96E-04 | 1.74E-03 | 2.42E-03 | 3.32E-03 | 2.72E-03 | 3.73E-04 | 2.3E-03 | 1.19E-03 | 2.41E-03 | 2.51E-03 | 2.66E-03 | 8.42E-04 | 3.87E-03 | 2.71E-03 | 3.21E-03 | 3.83E-03 | 3.13E-03 |
| ADENOSINE | 1.52E-02 | 2.87E-02 | 1.84E-02 | 2.51E-02 | 3.4E-02 | 8.26E-03 | 5.56E-03 | 1.07E-02 | 1.1E-02 | 1.4E-02 | 7.78E-03 | 4.33E-03 | 8.47E-03 | 1.83E-02 | 4.97E-03 | 2.26E-02 | 1.54E-02 | 1.58E-02 |
| ADENOSINE 5'-DIPHOSPHATE | 2.85E-04 | 3.87E-04 | 5.99E-04 | 2.07E-04 | 4.01E-04 | 3.22E-04 | 2.19E-04 | 2.11E-04 | 2.02E-04 | 2.2E-04 | 3.67E-04 | 4.77E-04 | 9.69E-04 | 6.84E-04 | 5.23E-04 | 7.22E-04 | 6.48E-04 | 5.44E-04 |
| ADENOSINE TRIPHOSPHATE | 8.92E-04 | 7.83E-04 | 9.7E-04 | 4.52E-04 | 1.11E-03 | 2.39E-04 | 1.78E-04 | 3.14E-04 | 4.3E-04 | 4.29E-04 | 6.45E-04 | 2.88E-03 | 1.21E-03 | 9.4E-04 | 2.55E-03 | 1.74E-03 | 3.32E-03 | 9.05E-04 |
| ALLANTOIN | 2.75E-04 | 3.93E-04 | 3.63E-04 | 2.99E-04 | 2.55E-04 | 2.89E-04 | 3.14E-04 | 2.76E-04 | 3.33E-04 | 3.07E-04 | 3.44E-04 | 2.37E-04 | 3.46E-04 | 2.27E-04 | 2.41E-04 | 2.24E-04 | 2.32E-04 | 2.83E-04 |
| AMINOISOBUTANOATE | 2.01E-05 | 5.68E-05 | 4.09E-05 | 3.33E-05 | 5.1E-05 | 2.39E-05 | 3.04E-05 | 1.87E-05 | 2.41E-05 | 3.5E-05 | 3.16E-05 | 3.93E-05 | 6.17E-05 | 3.79E-05 | 2.88E-05 | 3.96E-05 | 1.37E-05 | 4.22E-05 |
| ARABINOSE | 1.69E-04 | 2.63E-04 | 2.87E-04 | 1.56E-04 | 2.52E-04 | 1.54E-04 | 2.01E-04 | 1.33E-04 | 1.36E-04 | 2.72E-04 | 1.97E-04 | 1.98E-04 | 5.79E-04 | 2.42E-04 | 2.02E-04 | 3.92E-04 | 3.53E-04 | 3.16E-04 |
| ASPARAGINE | 1.7E-04 | 1.7E-04 | 2.6E-04 | 4E-05 | 6.86E-05 | 4.38E-05 | 4.12E-05 | 7.68E-07 | 9.56E-07 | 5.19E-05 | 6.12E-05 | 8.53E-07 | 1.53E-06 | 1.42E-04 | 1.08E-04 | 1.23E-06 | 7.48E-05 | 1.01E-06 |
| AZELATE | 1.27E-04 | 1.2E-04 | 1.58E-04 | 1.02E-04 | 1E-04 | 6.25E-05 | 1.26E-04 | 6.58E-05 | 8.05E-05 | 1.3E-04 | 1.21E-04 | 1.14E-04 | 4.16E-04 | 1.3E-04 | 7.21E-05 | 3.61E-04 | 1.33E-04 | 1.85E-04 |
| BENZOATE | 1.65E-02 | 2.54E-02 | 2.2E-02 | 1.57E-02 | 2.25E-02 | 1.73E-02 | 1.96E-02 | 1.72E-02 | 2.08E-02 | 2.19E-02 | 1.88E-02 | 1.67E-02 | 2.87E-02 | 1.99E-02 | 1.47E-02 | 2.35E-02 | 1.82E-02 | 2.01E-02 |
| BENZYL ALCOHOL | 1.05E-03 | 2.1E-03 | 1.65E-03 | 9.96E-04 | 1.57E-03 | 1.14E-03 | 1.32E-03 | 1.08E-03 | 1.29E-03 | 1.39E-03 | 2.99E-03 | 1.14E-03 | 2.3E-03 | 1.48E-03 | 1.06E-03 | 1.57E-03 | 1.25E-03 | 2.12E-03 |
| CAPRYLATE | 2.59E-04 | 2.71E-04 | 3.53E-04 | 2.66E-04 | 3.86E-04 | 2.62E-04 | 3.35E-04 | 2.98E-04 | 3.21E-04 | 2.52E-04 | 3.8E-04 | 2.66E-04 | 4.31E-04 | 3.03E-04 | 2.73E-04 | 3.45E-04 | 3.31E-04 | 3.51E-04 |
| CITRAMALATE | 4.75E-04 | 6.7E-04 | 7.12E-04 | 4.77E-04 | 5.17E-04 | 4.21E-04 | 4.53E-04 | 2.87E-04 | 4.06E-04 | 3.3E-04 | 4.84E-04 | 5.57E-04 | 9.58E-04 | 5.31E-04 | 4.45E-04 | 7.64E-04 | 6.38E-04 | 7.19E-04 |
| CYSTEATE | 1.89E-04 | 1.8E-04 | 3.28E-04 | 4.23E-05 | 3.74E-05 | 3.94E-05 | 5.8E-05 | 2.24E-05 | 2.11E-05 | 3.39E-05 | 4.74E-05 | 6.58E-05 | 1.93E-04 | 1.45E-04 | 1.65E-04 | 1.68E-04 | 1.58E-04 | 1.88E-04 |
| CYSTEINE | 2.84E-05 | 2.79E-05 | 6.77E-05 | 4.74E-05 | 4.12E-05 | 4.05E-05 | 9.02E-07 | 7.68E-07 | 2.89E-05 | 9.7E-07 | 3.28E-05 | 5.49E-05 | 1.53E-06 | 1.01E-06 | 7.04E-05 | 1.23E-06 | 9.77E-07 | 3.08E-05 |
| DECANOATE | 7.23E-06 | 1.42E-05 | 1.62E-05 | 9.87E-06 | 1.22E-05 | 8.11E-07 | 8.57E-06 | 1.02E-05 | 8.26E-06 | 1.15E-05 | 9.48E-06 | 1.22E-05 | 2.06E-05 | 1.78E-05 | 8.73E-06 | 1.12E-05 | 1.25E-05 | 1.82E-05 |
| DEOXYCARNITINE | 1.07E-03 | 1.02E-02 | 6.32E-03 | 3.08E-02 | 3.23E-02 | 2.08E-03 | 7.33E-03 | 3.96E-02 | 4.43E-02 | 4.97E-03 | 4.86E-02 | 3.02E-02 | 8.61E-03 | 4.88E-03 | 2.39E-03 | 6.94E-03 | 1.98E-03 | 1.1E-02 |
| DEOXYGUANOSINE DIPHOSPHATE | 2.03E-05 | 3.89E-05 | 3.22E-05 | 1.51E-05 | 2.33E-05 | 1.56E-05 | 9.02E-07 | 1.36E-05 | 9.56E-07 | 1.95E-05 | 1.59E-05 | 2.11E-05 | 1.53E-06 | 2.14E-05 | 2.35E-05 | 4.42E-05 | 3.01E-05 | 3.21E-05 |
| DL-2-AMINOCTANOATE | 1.35E-04 | 4.77E-04 | 1.72E-04 | 1.34E-04 | 2.68E-04 | 1.45E-04 | 2.6E-04 | 1.4E-04 | 1.85E-04 | 2.37E-04 | 8.75E-04 | 1.81E-04 | 3.88E-04 | 3.86E-04 | 1.69E-04 | 1.45E-04 | 1.2E-04 | 2.86E-04 |
| FOLATE | 3.28E-04 | 2.6E-04 | 7.59E-04 | 3.92E-04 | 3.56E-04 | 4.21E-04 | 2.38E-04 | 1.43E-04 | 2.08E-04 | 2.54E-04 | 2.55E-04 | 5.71E-04 | 1.08E-03 | 6.18E-04 | 5.94E-04 | 6.1E-04 | 5.08E-04 | 5.29E-04 |
| FRUCTOSE | 3.63E-04 | 4.56E-04 | 1.1E-06 | 3.89E-04 | 1.03E-06 | 4.22E-04 | 4.6E-04 | 3.73E-04 | 4.2E-04 | 3.61E-04 | 4.16E-04 | 3.46E-04 | 1.53E-06 | 3.49E-04 | 3.82E-04 | 1.23E-06 | 9.77E-07 | 4.39E-04 |
| FRUCTOSE 6-PHOSPHATE | 4.31E-04 | 5.06E-04 | 8.37E-04 | 3.86E-04 | 5.18E-04 | 3.78E-04 | 9.02E-07 | 1.94E-04 | 9.56E-07 | 2.83E-04 | 3.68E-04 | 4.62E-04 | 1.4E-03 | 9.65E-04 | 7.27E-04 | 5.33E-04 | 4.02E-04 | 4.18E-04 |
| FUCOSE | 1.37E-03 | 1.37E-03 | 3.91E-03 | 3.44E-04 | 2.34E-03 | 8.82E-04 | 1.81E-03 | 3.19E-05 | 4.78E-04 | 2.75E-03 | 3.28E-04 | 2.1E-03 | 4.59E-03 | 3.79E-04 | 1.24E-04 | 1.73E-03 | 1.47E-03 | 6.44E-04 |
| GALACTOSE | 6.35E-04 | 8.43E-04 | 1.21E-03 | 5.65E-04 | 8.22E-04 | 6.19E-04 | 7.32E-04 | 4.97E-04 | 5.89E-04 | 8.18E-04 | 6.52E-04 | 6.98E-04 | 1.79E-03 | 6.74E-04 | 5.49E-04 | 1.01E-03 | 8.63E-04 | 8.23E-04 |
| GALACTURONATE | 1.34E-04 | 1.67E-04 | 2.09E-04 | 1.09E-04 | 1.21E-04 | 1.03E-04 | 1.11E-04 | 9.34E-05 | 1.1E-04 | 1.08E-04 | 1.77E-04 | 1.57E-04 | 2.39E-04 | 1.01E-06 | 1.39E-04 | 1.23E-06 | 9.77E-07 | 1.88E-04 |
| GLUCONATE | 9.2E-04 | 1.13E-03 | 1.55E-03 | 4.25E-04 | 5.37E-04 | 1.62E-04 | 4.81E-04 | 3.32E-04 | 3.58E-04 | 4.36E-04 | 5.77E-04 | 7.03E-04 | 1.27E-03 | 6.33E-04 | 6.51E-04 | 6.58E-04 | 6.76E-04 | 6.49E-04 |
| GLUCOSE | 5.63E-04 | 7.59E-04 | 1.09E-03 | 4.69E-04 | 7.07E-04 | 5.19E-04 | 6.1E-04 | 4.43E-04 | 4.7E-04 | 6.75E-04 | 5.9E-04 | 6.41E-04 | 1.65E-03 | 6.09E-04 | 4.78E-04 | 9.59E-04 | 7.61E-04 | 7.49E-04 |
| GLUCOSE 1-PHOSPHATE | 7.29E-05 | 1.12E-04 | 1.8E-04 | 7.67E-05 | 1.06E-04 | 8.53E-05 | 5.85E-05 | 4.78E-05 | 5.45E-05 | 5.02E-05 | 5.1E-05 | 9.85E-05 | 2.99E-04 | 2.06E-04 | 1.4E-04 | 9.37E-05 | 5.23E-05 | 8.17E-05 |
| GLUCOSE 6-PHOSPHATE | 4.29E-04 | 4.99E-04 | 8.33E-04 | 3.84E-04 | 5.14E-04 | 8.11E-07 | 9.02E-07 | 1.95E-04 | 9.56E-07 | 2.86E-04 | 3.6E-04 | 4.59E-04 | 1.39E-03 | 9.78E-04 | 7.21E-04 | 5.28E-04 | 3.97E-04 | 4.09E-04 |
| GLUCURONATE | 3.91E-05 | 9.52E-05 | 2.64E-05 | 1.61E-05 | 4.39E-05 | 6.02E-05 | 7.74E-05 | 7.31E-05 | 5.28E-05 | 8.15E-05 | 1.43E-05 | 5.68E-05 | 2.94E-05 | 6.01E-05 | 2.49E-05 | 5.28E-05 | 5.8E-05 | 2.58E-05 |
| GLUTAMATE | 4.29E-02 | 5.4E-02 | 6.47E-02 | 2.97E-02 | 4.13E-02 | 2.67E-02 | 2.75E-02 | 1.79E-02 | 1.96E-02 | 2.88E-02 | 4.06E-02 | 4.08E-02 | 6.94E-02 | 5.18E-02 | 4.41E-02 | 6.4E-02 | 5.79E-02 | 5.58E-02 |
| GLYCEROPHOSPHOCHOLINE | 6.19E-05 | 1.66E-04 | 1.06E-04 | 6.19E-05 | 7.04E-05 | 2.85E-05 | 7.21E-05 | 1.67E-04 | 2.02E-04 | 2.5E-05 | 2.43E-04 | 8.53E-07 | 2.59E-04 | 6.83E-05 | 3.13E-05 | 2.12E-04 | 1.94E-05 | 1.56E-04 |
| GLYCINE | 3.06E-05 | 4.66E-05 | 1.6E-05 | 1.77E-05 | 2.97E-05 | 3.2E-05 | 3.29E-05 | 1.9E-05 | 2.54E-05 | 2.35E-05 | 3.46E-05 | 2.67E-05 | 4.11E-05 | 2.79E-05 | 1.79E-05 | 3.94E-05 | 3.38E-05 | 4.5E-05 |
| GLYCOCHENODEOXYCHOLATE | 1.6E-03 | 1.55E-03 | 1.98E-03 | 1.81E-03 | 1.03E-03 | 1.99E-03 | 2.02E-03 | 1.83E-03 | 1.53E-03 | 1.25E-03 | 1.27E-03 | 7.89E-04 | 1.45E-03 | 1.33E-03 | 7.64E-04 | 8.23E-04 | 1.21E-03 | 1.53E-03 |
| GUANINE-2 | 8.25E-07 | 1.79E-04 | 1.79E-04 | 9.08E-05 | 1.03E-06 | 1.14E-04 | 1.23E-04 | 9.84E-05 | 9.56E-07 | 9.7E-07 | 9.13E-07 | 1.2E-04 | 2.59E-04 | 1.26E-04 | 1.1E-04 | 1.38E-04 | 1.32E-04 | 1.68E-04 |
| GUANOSINE DIPHOSPHATE MANNOSE | 3.29E-05 | 3.89E-05 | 6.76E-05 | 3.86E-05 | 4.6E-05 | 2.4E-05 | 2.03E-05 | 1.65E-05 | 1.24E-05 | 1.53E-05 | 3.45E-05 | 2.63E-05 | 1.12E-04 | 7.52E-05 | 7.14E-05 | 7.91E-05 | 8.74E-05 | 6.05E-05 |
| HEXANOATE | 2.06E-04 | 2.67E-04 | 3.61E-04 | 1.56E-04 | 2.85E-04 | 1.7E-04 | 2.15E-04 | 1.74E-04 | 2.47E-04 | 2.75E-04 | 3.37E-04 | 2.17E-04 | 5.93E-04 | 2.8E-04 | 2.02E-04 | 3.39E-04 | 2.75E-04 | 3.63E-04 |
| HEXANOYLGLYCINE | 1.48E-04 | 2.31E-04 | 5.42E-04 | 1.46E-04 | 1.03E-06 | 2.33E-04 | 9.48E-05 | 7.68E-07 | 9.56E-07 | 9.7E-07 | 2.07E-04 | 3.11E-04 | 1.53E-06 | 7.36E-04 | 6.24E-04 | 1.23E-06 | 6.69E-04 | 3.99E-04 |
| HIPPURATE | 1.56E-03 | 2.28E-03 | 2.36E-03 | 1.71E-03 | 1.43E-03 | 1.86E-03 | 2.02E-03 | 1.59E-03 | 2.38E-03 | 1.79E-03 | 1.98E-03 | 1.45E-03 | 2.45E-03 | 1.21E-03 | 1.38E-03 | 1.25E-03 | 1.2E-03 | 1.41E-03 |
| HISTAMINE | 6.25E-05 | 1.82E-05 | 8.99E-05 | 3.35E-05 | 6.15E-05 | 5.29E-05 | 5E-05 | 1.25E-04 | 1.33E-05 | 5.06E-05 | 1.76E-04 | 5.17E-05 | 1.8E-04 | 1.15E-04 | 9.31E-05 | 9.15E-05 | 1.21E-04 | 1.48E-04 |
| HOMOSERINE | 7.33E-04 | 1.17E-03 | 1.08E-03 | 4.7E-04 | 6.83E-04 | 4.56E-04 | 5.4E-04 | 3.96E-04 | 4E-04 | 5.07E-04 | 7.45E-04 | 5.72E-04 | 1.36E-03 | 8.22E-04 | 6.47E-04 | 1.58E-03 | 1.27E-03 | 1.23E-03 |
| HOMOVANILLATE | 1.84E-04 | 2.66E-04 | 2.33E-04 | 1.86E-04 | 1.67E-04 | 1.87E-04 | 2.16E-04 | 1.68E-04 | 2.4E-04 | 1.99E-04 | 2.15E-04 | 1.73E-04 | 2.31E-04 | 1.32E-04 | 1.43E-04 | 1.37E-04 | 1.52E-04 | 1.57E-04 |
| HYDROXYPHENYLLACTATE | 1.84E-04 | 2.62E-04 | 2.33E-04 | 1.82E-04 | 1.68E-04 | 1.84E-04 | 2.16E-04 | 1.68E-04 | 2.39E-04 | 2.01E-04 | 2.12E-04 | 1.68E-04 | 2.29E-04 | 1.33E-04 | 1.43E-04 | 1.48E-04 | 1.5E-04 | 1.47E-04 |
| HYPOXANTHINE | 4.71E-03 | 8.55E-03 | 8.25E-03 | 4.35E-03 | 4.96E-03 | 4.88E-03 | 4.64E-03 | 4.55E-03 | 6.88E-03 | 3.94E-03 | 6.4E-03 | 5.37E-03 | 7.4E-03 | 6.01E-03 | 5.05E-03 | 4.39E-03 | 4.81E-03 | 6.73E-03 |
| ISOLEUCINE | 5.43E-04 | 9.97E-04 | 1.28E-03 | 7.45E-04 | 2.31E-03 | 1.04E-03 | 2.31E-03 | 8.79E-04 | 1.19E-03 | 1.85E-03 | 6.34E-04 | 1.77E-03 | 2.93E-03 | 1.29E-03 | 5.18E-04 | 1.45E-03 | 1.73E-03 | 6.11E-04 |
| ITACONATE | 1.03E-04 | 1.65E-04 | 1.69E-04 | 1.04E-04 | 1.44E-04 | 1.05E-04 | 1.05E-04 | 7.46E-05 | 9.01E-05 | 8.87E-05 | 1.26E-04 | 1.36E-04 | 2.87E-04 | 1.78E-04 | 1.45E-04 | 2.51E-04 | 1.76E-04 | 1.89E-04 |
| KETOLEUCINE | 8.23E-05 | 1.53E-04 | 1.72E-04 | 7.95E-05 | 1.19E-04 | 1.01E-04 | 1.15E-04 | 9.46E-05 | 1.12E-04 | 1.29E-04 | 1.2E-04 | 1.31E-04 | 2.23E-04 | 1.55E-04 | 1.05E-04 | 1.21E-04 | 1.14E-04 | 1.66E-04 |

| | | | | | | | | | | | | | | | | | | |
|---|---|---|---|---|---|---|---|---|---|---|---|---|---|---|---|---|---|---|
| **KYNURENINE** | 1.46E-04 | 2.74E-04 | 2.81E-04 | 2.21E-04 | 2.59E-04 | 2.39E-04 | 1.65E-04 | 1.52E-04 | 1.79E-04 | 2.14E-04 | 2.94E-04 | 1.71E-04 | 4.43E-04 | 2.66E-04 | 2.6E-04 | 1.23E-06 | 9.77E-07 | 1.01E-06 |
| **LEUCINE** | 1.5E-02 | 2.36E-02 | 2.01E-02 | 1.52E-02 | 1.62E-02 | 1.59E-02 | 1.82E-02 | 1.55E-02 | 2.08E-02 | 1.61E-02 | 1.86E-02 | 1.35E-02 | 2.27E-02 | 1.33E-02 | 1.4E-02 | 1.81E-02 | 1.35E-02 | 1.75E-02 |
| **MALEAMATE** | 2.81E-05 | 3.84E-05 | 3.23E-05 | 1.63E-05 | 1.62E-05 | 1.91E-05 | 1.36E-05 | 1.59E-05 | 1.9E-05 | 1.85E-05 | 2.62E-05 | 1.73E-05 | 2.51E-05 | 1.16E-05 | 1.5E-05 | 2.16E-05 | 1.53E-05 | 2.05E-05 |
| **METHIONINE** | 1.68E-05 | 1.27E-05 | 1.52E-05 | 1.78E-05 | 1.39E-05 | 1.63E-05 | 9.31E-06 | 2.06E-05 | 1.92E-05 | 1.49E-05 | 2.12E-05 | 2.49E-05 | 1.1E-05 | 3.89E-05 | 3.01E-05 | 1.98E-05 | 3.21E-05 | 2.16E-05 |
| **METHYL ACETOACETATE** | 2.21E-05 | 1.76E-05 | 2.48E-05 | 2.41E-05 | 5.57E-05 | 3.37E-05 | 4.2E-05 | 4.14E-05 | 2.9E-05 | 1.31E-05 | 2.13E-05 | 2.47E-05 | 6.18E-05 | 5.47E-05 | 3.54E-05 | 2.99E-05 | 3.13E-05 | 3.74E-05 |
| **METHYLMALONATE** | 2.06E-04 | 2.16E-04 | 3.36E-04 | 1.87E-04 | 1.71E-04 | 1.74E-04 | 1.87E-04 | 1.08E-04 | 1.65E-04 | 1.46E-04 | 2.03E-04 | 2.73E-04 | 3.83E-04 | 2.02E-04 | 2.26E-04 | 2.6E-04 | 2.31E-04 | 2.45E-04 |
| **MEVALOLACTONE** | 1.56E-04 | 2.4E-04 | 1.98E-04 | 1.32E-04 | 1.78E-04 | 1.64E-04 | 1.5E-04 | 1.6E-04 | 1.95E-04 | 1.48E-04 | 1.53E-04 | 1.21E-04 | 2.25E-04 | 1.34E-04 | 1.38E-04 | 1.83E-04 | 1.46E-04 | 1.58E-04 |
| **MYOINOSITOL** | 1.87E-03 | 2.51E-03 | 2.65E-03 | 9.36E-04 | 1.27E-03 | 9.53E-04 | 1.63E-03 | 1.11E-03 | 1.26E-03 | 1.67E-03 | 2.13E-03 | 1.94E-03 | 1.83E-03 | 9.5E-04 | 8.57E-04 | 2.49E-03 | 2.14E-03 | 2.09E-03 |
| **N-a-Acetyl-L-arginine** | 4.44E-03 | 6.73E-03 | 5.03E-03 | 4.66E-03 | 6.13E-03 | 4.25E-03 | 5.33E-03 | 4.32E-03 | 4.88E-03 | 4.73E-03 | 4.99E-03 | 5.41E-03 | 4.73E-03 | 5.84E-03 | 5.05E-03 | 5.8E-03 | 4.84E-03 | 5.59E-03 |
| **N-ACETYLASPARAGINE** | 1.98E-04 | 4.83E-04 | 2.5E-05 | 1.48E-04 | 2.06E-04 | 1.37E-04 | 1.81E-04 | 1.46E-04 | 2.03E-04 | 1.58E-04 | 2.49E-04 | 2.23E-04 | 3.81E-04 | 2E-04 | 2.21E-04 | 3.24E-04 | 2.85E-04 | 3.7E-05 |
| **N-ACETYLASPARTATE** | 3.06E-03 | 4.4E-03 | 4.67E-03 | 1.55E-03 | 2.07E-03 | 1.48E-03 | 1.46E-03 | 1.06E-03 | 1.24E-03 | 1.47E-03 | 2.36E-03 | 2.61E-03 | 3.37E-03 | 2.06E-03 | 1.71E-03 | 3.61E-03 | 3.16E-03 | 3.17E-03 |
| **N-ACETYLGLUTAMATE** | 1.67E-03 | 1.97E-03 | 2.43E-03 | 1.31E-03 | 1.46E-03 | 1.15E-03 | 1.55E-03 | 9.13E-04 | 9.83E-04 | 8.64E-04 | 1.69E-03 | 1.85E-03 | 4.28E-03 | 1.81E-03 | 1.78E-03 | 3.96E-03 | 3.13E-03 | 3.33E-03 |
| **N-ACETYLGLYCINE** | 8.83E-05 | 1.28E-04 | 1.1E-06 | 9.46E-05 | 1.03E-06 | 8.19E-05 | 1.04E-04 | 8.06E-05 | 8.71E-05 | 1E-04 | 1.54E-04 | 8.53E-07 | 1.53E-06 | 1.65E-04 | 1.31E-04 | 1.23E-06 | 9.77E-07 | 2.29E-04 |
| **N-ACETYLLEUCINE** | 1.75E-04 | 2.04E-04 | 2.56E-04 | 1.71E-04 | 2E-04 | 2E-04 | 1.84E-04 | 1.59E-04 | 3E-04 | 2.07E-04 | 2.77E-04 | 2.01E-04 | 3.81E-04 | 1.19E-04 | 1.59E-04 | 1.41E-04 | 1.32E-04 | 1.58E-04 |
| **N-ACETYLMETHIONINE** | 1.01E-03 | 1.65E-03 | 1.43E-03 | 5.52E-04 | 8.12E-04 | 5.85E-04 | 8.21E-04 | 6.36E-04 | 7.41E-04 | 8.56E-04 | 1.14E-03 | 8.03E-04 | 1.42E-03 | 6.59E-04 | 5.9E-04 | 1.29E-03 | 1.26E-03 | 1.11E-03 |
| **N-ACETYLNEURAMINATE** | 1.46E-03 | 1.2E-06 | 1.1E-06 | 1.42E-03 | 2.01E-03 | 1.38E-03 | 1.63E-03 | 7.68E-07 | 9.56E-07 | 1.2E-03 | 9.13E-07 | 1.89E-03 | 1.53E-06 | 1.59E-03 | 1.4E-03 | 1.23E-06 | 2.18E-03 | 1.01E-06 |
| **N-ACETYLPHENYLALANINE** | 2.81E-04 | 6.45E-04 | 4.94E-04 | 3.41E-04 | 4.54E-04 | 3.06E-04 | 3.33E-04 | 3.55E-04 | 5.43E-04 | 4.27E-04 | 5.31E-04 | 3.88E-04 | 5.02E-04 | 2.21E-04 | 3.17E-04 | 9.49E-05 | 2.23E-04 | 2.42E-04 |
| **N-ACETYLSERINE** | 6.59E-04 | 9.41E-04 | 1.1E-06 | 6.1E-04 | 7.76E-04 | 5.35E-04 | 7.14E-04 | 5.82E-04 | 5.8E-04 | 7.2E-04 | 1.04E-03 | 8.53E-07 | 1.53E-06 | 1.06E-03 | 9.21E-04 | 1.23E-06 | 9.77E-07 | 1.56E-03 |
| **N-FORMYLMETHIONINE** | 1.18E-04 | 2.58E-04 | 1.9E-04 | 6.98E-05 | 9.78E-05 | 7.89E-05 | 7.13E-05 | 7.15E-05 | 7.89E-05 | 7.22E-05 | 8.46E-05 | 7.83E-05 | 1.24E-04 | 7.31E-05 | 7.04E-05 | 1.32E-04 | 1.2E-04 | 9.39E-05 |
| **NICOTINAMIDE** | 3.43E-02 | 1.5E-02 | 1.18E-02 | 7.36E-05 | 1.46E-02 | 3.92E-05 | 2.73E-02 | 3.48E-02 | 2.66E-02 | 2.66E-02 | 2.24E-02 | 2.26E-02 | 6.84E-03 | 1.92E-02 | 2.63E-02 | 1.36E-02 | 1.92E-02 | 1.61E-02 |
| **NORADRENALINE** | 1.32E-01 | 8.98E-02 | 8.38E-02 | 1.28E-01 | 1.09E-01 | 1.39E-01 | 1.28E-01 | 1.38E-01 | 1.2E-01 | 1.33E-01 | 1.02E-01 | 1.18E-01 | 4.49E-02 | 1.03E-01 | 1.18E-01 | 9.06E-02 | 1.03E-01 | 9.71E-02 |
| **NORVALINE** | 4.79E-03 | 6.75E-03 | 6.98E-03 | 4.74E-03 | 4.92E-03 | 4.93E-03 | 5.48E-03 | 3.9E-03 | 5.32E-03 | 4.62E-03 | 5.65E-03 | 4.36E-03 | 6.94E-03 | 3.85E-03 | 4.18E-03 | 4.69E-03 | 4.54E-03 | 4.93E-03 |
| **OROTATE** | 1.77E-05 | 3.06E-05 | 3.18E-05 | 1.51E-05 | 2.96E-05 | 2.3E-05 | 2.16E-05 | 1.52E-05 | 2.15E-05 | 2.86E-05 | 2.21E-05 | 2.29E-05 | 4.43E-05 | 2.81E-05 | 1.87E-05 | 2.57E-05 | 2.54E-05 | 2.23E-05 |
| **OXOADIPATE** | 6.49E-05 | 6.88E-05 | 7.67E-05 | 4.33E-05 | 6.45E-05 | 4.7E-05 | 5.78E-05 | 7.68E-07 | 9.56E-07 | 4.72E-05 | 5.97E-05 | 6.6E-05 | 1.44E-04 | 7.39E-05 | 5.43E-05 | 9.07E-05 | 5.81E-05 | 6.36E-05 |
| **OXOPROLINE** | 7.57E-02 | 9.11E-02 | 1.14E-01 | 7.56E-02 | 7.25E-02 | 8.26E-02 | 9.06E-02 | 6.32E-02 | 8.43E-02 | 7.78E-02 | 8.28E-02 | 7.95E-02 | 1.21E-01 | 7.03E-02 | 7.44E-02 | 7.1E-02 | 6.94E-02 | 7.73E-02 |
| **PHENOL** | 1.73E-03 | 2.87E-03 | 2.3E-03 | 1.54E-03 | 2.14E-03 | 1.67E-03 | 1.95E-03 | 1.72E-03 | 2.38E-03 | 2.52E-03 | 2.03E-03 | 2.06E-03 | 2.92E-03 | 1.95E-03 | 1.43E-03 | 2.37E-03 | 1.8E-03 | 2.08E-03 |
| **PHENYLALANINE** | 7.33E-02 | 1.1E-01 | 9.33E-02 | 7.44E-02 | 7.97E-02 | 7.09E-02 | 7.96E-02 | 7.16E-02 | 9.19E-02 | 7.82E-02 | 9.87E-02 | 6.67E-02 | 1.05E-01 | 6.23E-02 | 6.03E-02 | 7.26E-02 | 6.95E-02 | 7.55E-02 |
| **PHOSPHORYLCHOLINE** | 8.25E-07 | 5.96E-02 | 4.5E-02 | 3.49E-02 | 3.76E-02 | 3.14E-02 | 3.81E-02 | 3.87E-02 | 4.21E-02 | 3.52E-02 | 6.12E-02 | 3.35E-02 | 5.66E-02 | 5.42E-02 | 3.82E-02 | 4.09E-02 | 2.99E-02 | 5.72E-02 |
| **PROPIONYLCARNITINE** | 1.29E-02 | 2.11E-02 | 1.98E-02 | 1.2E-02 | 1.51E-02 | 1.34E-02 | 6.81E-03 | 6.88E-03 | 8.27E-03 | 1.47E-02 | 1.86E-02 | 1.55E-02 | 2.84E-02 | 1.7E-02 | 1.64E-02 | 2.08E-02 | 1.97E-02 | 2.01E-02 |
| **PROPIONYLCHOLINE** | 1.12E-04 | 1.57E-04 | 8.48E-05 | 5.88E-05 | 6.43E-05 | 8.34E-05 | 8.69E-05 | 7.6E-05 | 4.58E-05 | 2.6E-05 | 2.19E-04 | 9.69E-06 | 3.13E-04 | 7E-05 | 8.24E-05 | 1.23E-06 | 9.14E-05 | 5.69E-05 |
| **PYRIDOXINE** | 1.31E-01 | 8.93E-02 | 8.26E-02 | 1.27E-01 | 1.08E-01 | 1.38E-01 | 1.28E-01 | 1.37E-01 | 1.19E-01 | 1.31E-01 | 1.02E-01 | 1.17E-01 | 4.48E-02 | 1.02E-01 | 1.16E-01 | 8.92E-02 | 1.03E-01 | 9.65E-02 |
| **RIBITOL** | 2.83E-05 | 5.98E-05 | 1.14E-04 | 5.93E-05 | 8.06E-05 | 8.11E-07 | 7.74E-05 | 8.24E-05 | 9.56E-07 | 7.92E-05 | 9.13E-07 | 7.44E-05 | 1.42E-04 | 9.38E-05 | 7.72E-07 | 1.24E-04 | 9.77E-07 | 1.01E-06 |
| **SALSOLINOL** | 8.25E-07 | 3.45E-03 | 1.1E-06 | 2.91E-04 | 5.98E-04 | 4.68E-04 | 5.24E-04 | 4.92E-04 | 9.56E-07 | 9.7E-07 | 5.87E-04 | 5.36E-04 | 3.18E-03 | 6.67E-04 | 3.4E-04 | 1.18E-03 | 7.11E-04 | 1.01E-06 |
| **SEBACATE** | 4.34E-05 | 6.23E-05 | 8.86E-05 | 4.79E-05 | 1.03E-06 | 5.04E-05 | 5.85E-05 | 4.34E-05 | 4.86E-05 | 6.58E-05 | 6.6E-05 | 8.53E-07 | 1.53E-06 | 6.46E-05 | 4.8E-05 | 1.04E-04 | 9.77E-07 | 8.06E-05 |
| **STEAROYLCARNITINE** | 8.25E-07 | 1.03E-04 | 7.03E-05 | 7.46E-07 | 1.03E-06 | 4.94E-05 | 3.57E-05 | 4.31E-05 | 9.56E-07 | 9.7E-07 | 1.27E-04 | 5.89E-05 | 7.65E-05 | 5.89E-05 | 3.93E-05 | 1.23E-06 | 9.77E-07 | 1.01E-06 |
| **SUBERATE** | 3.39E-05 | 3.43E-05 | 4.94E-05 | 2.44E-05 | 3.42E-05 | 1.98E-05 | 3.09E-05 | 2.32E-05 | 2.28E-05 | 3.55E-05 | 3.87E-05 | 2.94E-05 | 1.37E-04 | 6.79E-05 | 2.98E-05 | 8.7E-05 | 3.34E-05 | 6.02E-05 |
| **TARTRATE** | 6.11E-05 | 7.12E-05 | 1.11E-04 | 4.89E-05 | 6.83E-05 | 6.27E-05 | 5.2E-05 | 7.68E-07 | 4.6E-05 | 5.07E-05 | 6.82E-05 | 8.42E-05 | 2.05E-04 | 1.24E-04 | 1.25E-04 | 1.37E-04 | 7.54E-05 | 9.82E-05 |
| **TAURINE** | 1.28E-02 | 1.45E-02 | 2.16E-02 | 1.13E-02 | 1.76E-02 | 1.22E-02 | 1.52E-02 | 9.7E-03 | 9.27E-03 | 7.56E-03 | 1.02E-02 | 8.95E-03 | 4.12E-02 | 3.09E-02 | 2.78E-02 | 2.9E-02 | 2.7E-02 | 2.98E-02 |
| **THIOPURINE S-METHYLETHER** | 6.43E-05 | 8.52E-05 | 9.48E-05 | 5.36E-05 | 7.14E-05 | 4.98E-05 | 6.15E-05 | 6.01E-05 | 6.07E-05 | 5.3E-05 | 6.44E-05 | 5.77E-05 | 1.32E-04 | 8.39E-05 | 6.45E-05 | 1.01E-04 | 6.61E-05 | 7.49E-05 |
| **THYMIDINE** | 2.36E-04 | 1.2E-06 | 4.41E-04 | 1.94E-04 | 3.12E-04 | 3.61E-04 | 2.57E-04 | 7.68E-07 | 2.93E-04 | 4.71E-04 | 2.1E-04 | 8.53E-07 | 1.53E-06 | 2.81E-04 | 7.72E-07 | 3.97E-04 | 9.77E-07 | 1.01E-06 |
| **THYMINE** | 1.73E-04 | 3.06E-04 | 2.93E-04 | 1.74E-04 | 2.08E-04 | 1.87E-04 | 2.44E-04 | 1.89E-04 | 3.23E-04 | 2.56E-04 | 1.94E-04 | 1.85E-04 | 3.13E-04 | 1.67E-04 | 1.37E-04 | 1.11E-04 | 1.71E-04 | 1.76E-04 |
| **THYROXINE** | 9.99E-06 | 1.74E-05 | 2.33E-05 | 8.52E-06 | 1.85E-05 | 1.98E-05 | 1.85E-05 | 2.24E-05 | 2.98E-05 | 2.24E-05 | 1.06E-05 | 1.8E-05 | 1.98E-05 | 1.14E-05 | 1.26E-05 | 5.95E-06 | 1.08E-05 | 1.48E-05 |
| **TRANS-ACONITATE** | 6.75E-04 | 6.51E-04 | 1.16E-03 | 8.67E-04 | 9.6E-04 | 7.4E-04 | 5.17E-04 | 3.64E-04 | 4.99E-04 | 5.65E-04 | 7.28E-04 | 1.23E-03 | 2.6E-03 | 1.73E-03 | 1.41E-03 | 1.93E-03 | 1.6E-03 | 1.32E-03 |
| **TRANS-CYCLOHEXANEDIOL** | 2.06E-04 | 2.67E-04 | 3.61E-04 | 1.56E-04 | 2.86E-04 | 1.8E-04 | 2.15E-04 | 1.74E-04 | 2.28E-04 | 2.26E-04 | 3.71E-04 | 2.17E-04 | 5.93E-04 | 2.8E-04 | 2.03E-04 | 3.42E-04 | 2.75E-04 | 3.63E-04 |
| **UDP-GLUCOSE** | 2.75E-03 | 2.15E-03 | 5.19E-03 | 1.37E-03 | 1.73E-03 | 8.9E-04 | 7.59E-04 | 4.97E-04 | 1.18E-03 | 1.76E-03 | 1.31E-03 | 6.88E-03 | 8.41E-03 | 5.98E-03 | 6.31E-03 | 6.51E-03 | 7.18E-03 | 4.21E-03 |
| **UDP-N-ACETYLGLUCOSAMINE** | 2.91E-03 | 3.17E-03 | 6.74E-03 | 1.48E-03 | 2.07E-03 | 1.09E-03 | 7.96E-04 | 5.66E-04 | 6.28E-04 | 8.42E-04 | 1.81E-03 | 5.6E-03 | 9.07E-03 | 5.75E-03 | 6.19E-03 | 8.29E-03 | 7.31E-03 | 6.2E-03 |
| **URACIL** | 2.27E-04 | 3.78E-04 | 3.5E-04 | 2.43E-04 | 2.23E-04 | 3.01E-04 | 3.24E-04 | 2.59E-04 | 3.62E-04 | 2.81E-04 | 3.3E-04 | 2.02E-04 | 3.91E-04 | 1.81E-04 | 2.09E-04 | 2.11E-04 | 2.16E-04 | 2.43E-04 |
| **URACIL 5-CARBOXYLATE** | 8.25E-07 | 1.2E-06 | 1.1E-06 | 7.46E-07 | 7.96E-05 | 5.52E-05 | 5.92E-05 | 4.21E-05 | 9.56E-07 | 6.28E-05 | 9.13E-07 | 8.62E-05 | 2.17E-04 | 1.09E-04 | 4.52E-05 | 1.03E-04 | 8.98E-05 | 6.3E-05 |
| **URATE** | 3.27E-03 | 4.86E-03 | 4.58E-03 | 3.13E-03 | 3.54E-03 | 3.35E-03 | 3.81E-03 | 2.99E-03 | 3.65E-03 | 2.98E-03 | 4.1E-03 | 3.49E-03 | 5.5E-03 | 3.11E-03 | 2.97E-03 | 3.16E-03 | 2.82E-03 | 3.39E-03 |
| **URIDINE** | 5.38E-05 | 7.98E-05 | 7.74E-05 | 9.65E-05 | 1.08E-04 | 4.43E-05 | 4.09E-05 | 5.5E-05 | 8.54E-05 | 6.4E-05 | 2.96E-05 | 3.87E-05 | 8.43E-05 | 7.54E-05 | 3.29E-05 | 1.24E-04 | 8.07E-05 | 4.88E-05 |
| **URIDINE DIPHOSPHATE** | 1.27E-04 | 1.57E-04 | 2.75E-04 | 5.9E-05 | 9.95E-05 | 4.42E-05 | 3.83E-05 | 3.86E-05 | 9.56E-07 | 4.83E-05 | 8.92E-05 | 1.49E-04 | 3.07E-04 | 2.2E-04 | 3.09E-04 | 2.82E-04 | 3.47E-04 | 2.18E-04 |
| **URIDINE DIPHOSPHATE GLUCURONIC A** | 1.36E-04 | 2.33E-04 | 5.89E-04 | 7.43E-05 | 3.26E-04 | 6.62E-05 | 7.11E-05 | 1.12E-04 | 1.21E-04 | 1.47E-04 | 1.7E-04 | 2.9E-04 | 5.41E-04 | 3.52E-04 | 6.23E-04 | 3.46E-04 | 9.14E-04 | 5.84E-04 |
| **UROCANATE** | 8.25E-07 | 5.53E-04 | 8.35E-04 | 4.57E-04 | 1.03E-06 | 4.54E-04 | 5.52E-04 | 3.37E-04 | 9.56E-07 | 9.7E-07 | 4.7E-04 | 4.49E-04 | 1.53E-06 | 1.01E-06 | 3.54E-04 | 1.09E-03 | 4.92E-04 | 1.01E-06 |
| **VALINE** | 4.81E-03 | 6.75E-03 | 6.98E-03 | 4.75E-03 | 4.92E-03 | 4.92E-03 | 5.48E-03 | 3.89E-03 | 5.32E-03 | 4.62E-03 | 5.66E-03 | 4.36E-03 | 6.96E-03 | 3.84E-03 | 4.18E-03 | 4.69E-03 | 4.54E-03 | 4.92E-03 |
| **XANTHINE** | 1.46E-03 | 2.21E-03 | 2.5E-03 | 1.59E-03 | 1.38E-03 | 1.64E-03 | 1.91E-03 | 1.6E-03 | 1.95E-03 | 1.62E-03 | 2.02E-03 | 1.68E-03 | 2.23E-03 | 1.22E-03 | 1.47E-03 | 1.29E-03 | 1.31E-03 | 1.62E-03 |
